# Forecasting COVID-19 cases in the Philippines using various mathematical models

**DOI:** 10.1101/2020.10.07.20208421

**Authors:** Monica C. Torres, Christian Alvin H. Buhat, Ben Paul B. Dela Cruz, Edd Francis O. Felix, Eleanor B. Gemida, Jonathan B. Mamplata

## Abstract

Due to the rapid increase of COVID-19 infection cases in many countries such as the Philippines, many efforts in forecasting the daily infections have been made in order to better manage the pandemic, and respond effectively. In this study, we consider the cumulative COVID-19 infection cases in the Philippines from March 6 to July 31,2020 and forecast the cases from August 1 - 15, 2020 using various mathematical models —weighted moving average, exponential smoothing, Susceptible-Exposed-Infected-Recovered (SEIR) model, Ornstein-Uhlenbeck process, Autoregressive Integrated Moving Average (ARIMA) model, and random forest. We then compare the results to the actual data using traditional error metrics. Our results show that the ARIMA(1,2,1) model has the closest forecast values to the actual data. Policymakers can use our result in determining which forecast method to use for their community in order to have a data-based information for the preparation of their personnel and facilities.

## 1. Introduction

On December 31, 2019, the World Health Organization (WHO) was informed about an outbreak of pneumonia of an unknown cause in Wuhan City, Hubei Province of China [40]. Barely two weeks past, the first case of coronavirus disease 2019 (COVID-19) outside China was reported in a Chinese tourist in Thailand with no epidemiological linkage to the Huanan Seafood Wholesale Market [24]. Since then, COVID-19 quickly escalated worldwide infecting more than 200 countries and millions of people [41]. The COVID-19 which is caused by severe acute respiratory syndrome coronavirus 2 (SARS-CoV-2), is a kind of viral penumonia [28]. The most common symptoms at the onset of the disease include fever, cough, fatigue, dyspnea, myalgia, normal or decreased leukocyte counts, and radiographic evidence of pneumonia [28, 35]. Several evidences suggest that it can be transferred from person-to-person through direct contact or through droplets spread by coughing or sneezing from an infected individual [35].

Rapid increase in daily infection rates, severity of the cases, and death toll proved that the disease is highly contagious and infectious. In the absence of a vaccine, prevention and control are vital to suppress the spread of the virus in a way that the healthcare system can contain the surge of patients, and the economy can recuperate the impacts of lockdown restrictions. To this aim, modeling and forecasting are very important to lay down effective and data-based decisions, strategies, and policies. Since the outbreak of COVID-19, there have been many publications using various mathematical and machine learning-based models that forecast the spread and give a prediction of the epidemic peak globally, for specific countries such as in Brazil [34], Russia, India, and Bangladesh [32], and USA [2].

The dynamics of the COVID-19 infection vary per country and depend on many factors such as the government’s response, the capacity of the healthcare system, testing and contact tracing efforts, quarantine and lockdown impositions, and reaction of the general public to the pandemic. In the Philippines, the first case of COVID-19 was reported on January 30, 2020 while the first local transmission was confirmed on March 7, 2020 [39]. On March 17, 2020, the Philippine government declared an enhanced community quarantine (ECQ) in the entire island of Luzon and other parts of the county for a month and later extended until April 30 to curb the transmission of the disease [21]. Months since the lockdown, the economy gradually reopened with less stringent quarantine regulations in each province or region. However, the daily infection cases have not been significantly reduced yet. As of September 7, 2020, the total cases in the Philippines escalated to 238,727 [23]. Therefore, continuous effort to model and forecast the spread of the disease in the country is very helpful in the fight against COVID-19.

The UP-OCTA team has been actively forecasting the spread of the disease in different places in the Philippines. From April 30 to July 23, they were able to produce nine forecasts of COVID-19 cases using Susceptible-Exposed-Infected-Recovered (SEIR) models and Moving Average models [9, 8, 12, 13, 11, 7, 10, 15, 14]. The heterogeneous characteristics of different age populations were incorporated in studying the effects of the ECQ in reducing the exponential growth of the disease as well as the forecasts of transmission rate [17]. An SEIR model considering the symptomatic and asymptomatic populations was developed in [1] to describe the dynamics of the disease. The group used the data on the confirmed cases and death from several countries including France, Philippines, Italy, Spain, United Kingdom, China, and the USA to calibrate the model.

This study forecasts the cumulative daily cases of COVID-19 in the Philippines using various mathematical models. We examine six models and determine which best suits the considered Philippine data. The models under comparison are moving averages, exponential smoothing, SEIR, Ornstein-Uhlenbeck process, Autoregressive Integrated Moving Average (ARIMA), and random forest. The data set is fit using each of the models and subsequently, obtain forecasts for August 1-15. These forecasts are then compared to the actual values using various error metrics. The paper is organized as follows.

In Section 2, a description of the utilized data set is provided. Sections 3 to 8 discuss the implementation of the models and the derivation of the forecast values. The discussion on the comparison of the results from the six models is done in Sections 9 and 10.

## 2. Data Framework

Since April 12, COVID-19 data in the country have been made available through the COVID-19 tracker in order to promote transparency and accountability of data in the country. The tracker provides a daily COVID-19 data drop which contains multiple COVID-19 infection-related data based on multiple cases such as specimen collection, release of result, date of the start of symptom for each patient, date of reporting/ confirmation of positive COVID-19 result, etc. We only consider the infections based on the date of reporting/ confirmation of positive COVID-19 result, as this is the commonly reported data to the public. Shown in Table 1 are the descriptive statistics of the data to be considered and Figure 1 illustrates the actual March 6 to July 31 data.

**Table 1:** Descriptive statistics of the March 6 - July 31 COVID-19 data from the DOH data drop.

|  |  |
| --- | --- |
| Count | 148 |
| Minimum | 2 |
| Maximum | 93351 |
| Mean | 22328.60135 |
| Median | 12791.5 |
| Variance | 563842143.3 |
| Standard Deviation | 23825.9901 |
| Skewness | 1.280255329 |
| Kurtosis | 0.70664986 |
| Standard Error | 1958.484322 |
| Coefficient of Variation | 1.067061466 |

**Table 2:** 15-day ahead forecasts on the cumulative COVID-19 cases in the Philippines using 3-day, 4-day, 7-day, and 10-day SMA.

| Date | 3-day SMA | 4-day SMA | 7-day SMA | 10-day SMA |
| --- | --- | --- | --- | --- |
| 08/01/2020 | 96632 | 96219 | 95806 | 95648 |
| 08/02/2020 | 100384 | 99396 | 98328 | 98022 |
| 08/03/2020 | 104083 | 102902 | 100913 | 100415 |
| 08/04/2020 | 107660 | 106305 | 103632 | 102841 |
| 08/05/2020 | 111336 | 109543 | 106507 | 105309 |
| 08/06/2020 | 114986 | 112875 | 109525 | 107817 |
| 08/07/2020 | 118621 | 116244 | 112416 | 110410 |
| 08/08/2020 | 122275 | 119580 | 115140 | 113100 |
| 08/09/2020 | 125921 | 122898 | 117902 | 115873 |
| 08/10/2020 | 129566 | 126237 | 120698 | 118531 |
| 08/11/2020 | 133214 | 129578 | 123525 | 121049 |
| 08/12/2020 | 136861 | 132911 | 126367 | 123590 |
| 08/13/2020 | 140507 | 136244 | 129204 | 126146 |
| 08/14/2020 | 144154 | 139581 | 132015 | 128719 |
| 08/15/2020 | 147801 | 142917 | 134815 | 131307 |

**Table 3:** Weights used for the 4-day WMA.

| 4-day WMA | $r = 1$ | $r = 2$ | $r = 4$ | $r = 10$ |
| --- | --- | --- | --- | --- |
| $w_r(1)$ | 10% | 6.25% | 3.57% | 1.56% |
| $w_r(2)$ | 20% | 18.75% | 17.86% | 17.19% |
| $w_r(3)$ | 30% | 31.25% | 32.14% | 32.81% |
| $w_r(4)$ | 40% | 43.75% | 46.43% | 48.44% |

**Table 4:** 15-day ahead forecasts on the cumulative COVID-19 cases in the Philippines using 3-day, 4-day, 7-day, and 10-day WMA.

| Date | 3-day WMA | 4-day WMA | 7-day WMA | 10-day WMA |
| --- | --- | --- | --- | --- |
| 08/01/2020 | 97297 | 96949 | 96272 | 96000 |
| 08/02/2020 | 101281 | 100727 | 99305 | 98704 |
| 08/03/2020 | 105255 | 104497 | 102441 | 101461 |
| 08/04/2020 | 109231 | 108244 | 105648 | 104269 |
| 08/05/2020 | 113207 | 112002 | 108878 | 107122 |
| 08/06/2020 | 117183 | 115758 | 112072 | 110013 |
| 08/07/2020 | 121159 | 119514 | 115243 | 112922 |
| 08/08/2020 | 125135 | 123270 | 118424 | 115828 |
| 08/09/2020 | 129111 | 127026 | 121611 | 118705 |
| 08/10/2020 | 133087 | 130782 | 124800 | 121567 |
| 08/11/2020 | 137063 | 134539 | 127987 | 124434 |
| 08/12/2020 | 141038 | 138295 | 131173 | 127307 |
| 08/13/2020 | 145014 | 142051 | 134359 | 130183 |
| 08/14/2020 | 148990 | 145807 | 137545 | 133061 |
| 08/15/2020 | 152966 | 149563 | 140732 | 135938 |

**Figure 1:**
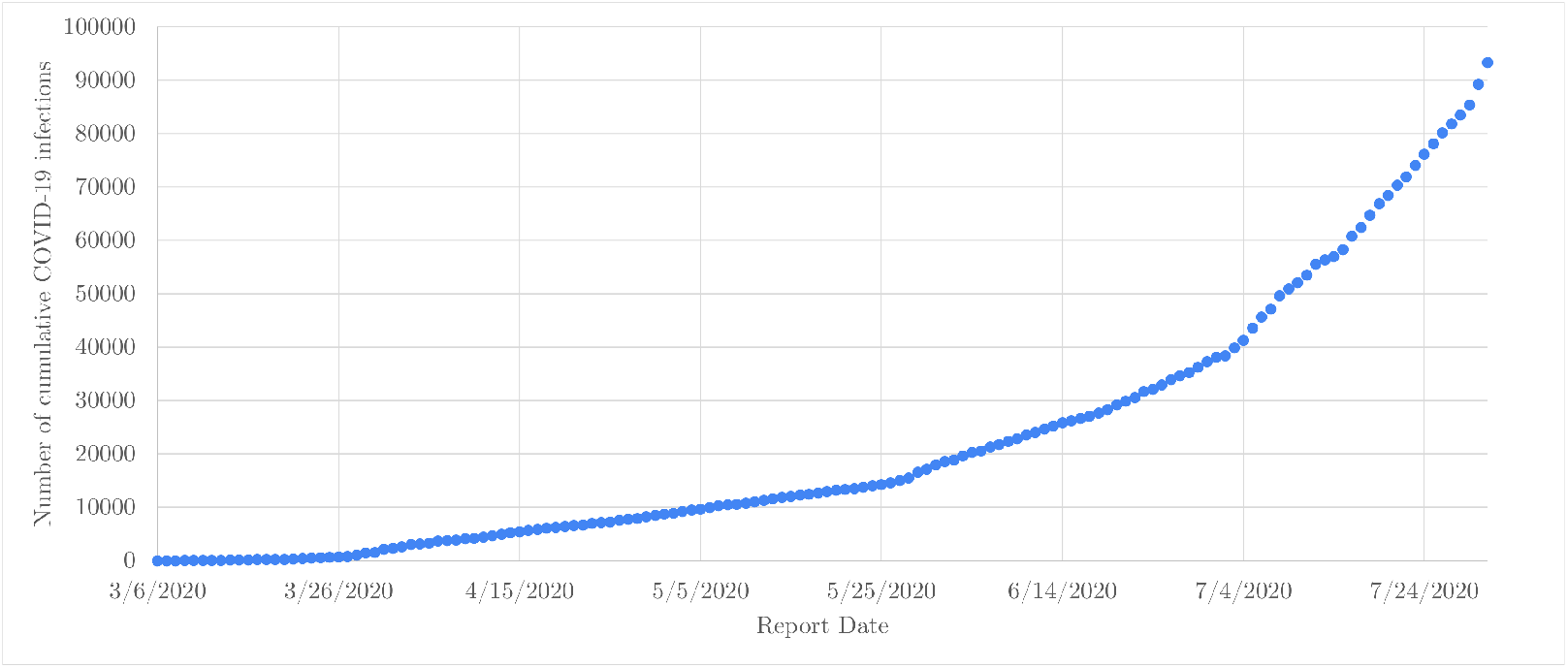
Actual cumulative COVID-19 cases of infection in the Philippines from March 6 - July 31, 2020.

**Figure 2:**
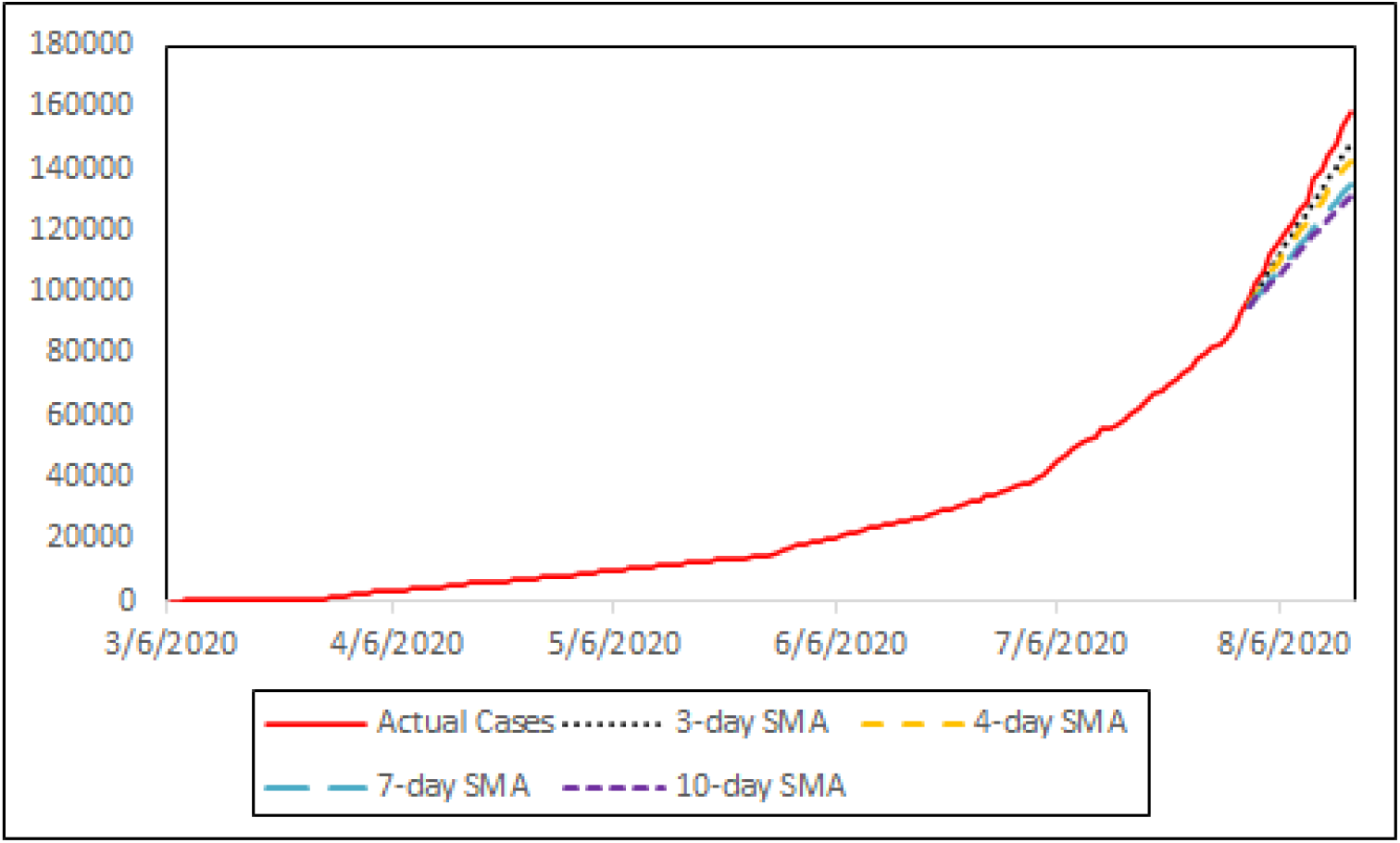
SMA forecast vs the actual data.

**Figure 3:**
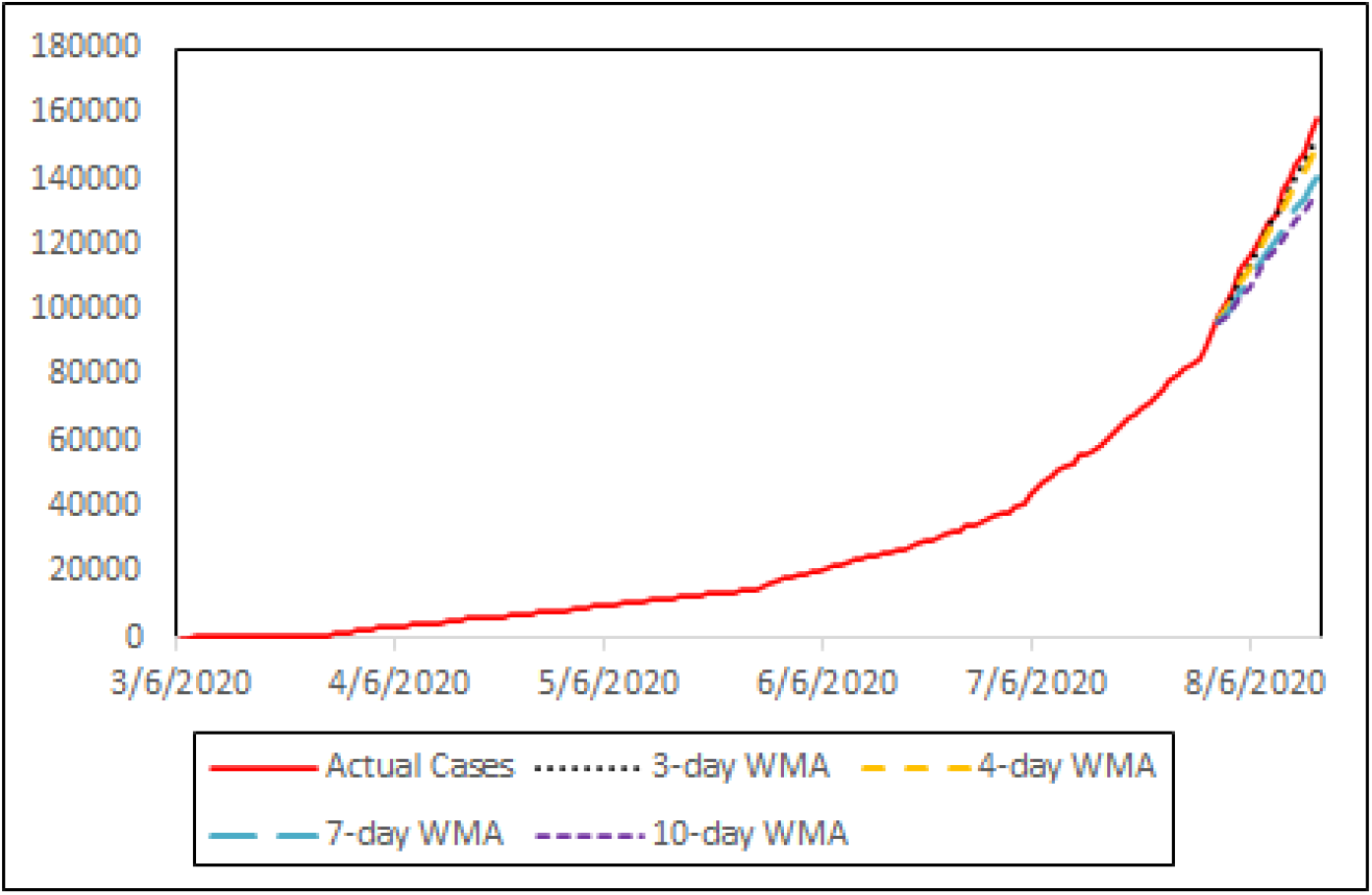
WMA with *r* = 10 forecast vs the actual data.

In this study, we consider the number of cumulative COVID-19 infection cases in the Philippines from March 6, 2020 to July 31, 2020 (retrieved: August 1, 2020). We then forecast the cumulative cases for August 1 - 15, and compare it to the actual values which were retrieved on August 24, 2020. We consider a half-month forecast, to account for the two-week quarantine period, to better prepare the concerned facilities/personnel, and to adjust accordingly to possible delays in reporting of cases.

## 3. Simple and Weighted Moving Averages

### 3.1. Preliminaries

In this section, we consider two types of moving average (MA) –the simple moving average (SMA) and the weighted moving average (WMA). We apply them to the cumulative cases to give forecasts for August 1 - 15. There are many different intervals used in literature regarding COVID-19 such as 3−day, 5−day, 7−day, 10−day, and 14 day [19, 22, 38]. In most cases, the 7−day MA is considered to cover both the incubation period and the time it takes from the first symptoms to occur to diagnosis [22].

The standard formula to get the forecast for the *n*^*th*^ day for an *m*-day SMA, is given by

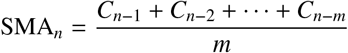

where *C*_*i*_ is the number of total confirmed cases at time *i*.

Simple moving average gives equal weights for all the data in a specific interval. While this is good for data sets whose entries are not dependent on the previous entries, it is far from good for those whose entries are highly dependent on the previous entries. Due to the increasing trend in the data set, it is better to consider higher weights in more recent entries. For this purpose, we also consider weighted moving average.

One of the most common weight functions used in weighted moving average is 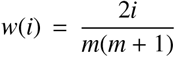 for the *i*^*th*^ day in the *m*-day interval. The standard formula to get the forecast for the *n*^*th*^ day for an *m*-day WMA is given by

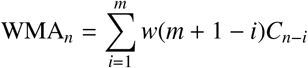

where *C*_*i*_ is the number of total confirmed cases at time *i*. Note that the sum of the weights in a weighted moving average must be equal to 1.

### 3.2. Numerical Implementation

We consider 3−day, 4−day, 7−day, and 10−day MA to have a comparison among different intervals for both types of moving average.

#### 3.2.1. Simple Moving Average

We apply SMA to the cumulative cases but the result was undesirable. For example, the actual cumulative cases on July 31 is 93351 while the forecast total number of COVID-19 cases on August 1 using 4-day SMA is 87881, which is inconsistent since the total cumulative cases must be increasing. In fact, the forecast for August 15 is 89556, which is still lower than the total cases on July 31. Instead of applying SMA directly to the cumulative cases, we applied SMA to the daily cases. To get the forecast for the cumulative cases for August 1, we add the forecast for the daily cases on August 1 to the actual total cases as of July 31.

Below is the table showing the forecasts using SMA on the different intervals mentioned.

#### 3.2.2. Weighted Moving Average

Similar to SMA, we apply WMA to the daily cases then we add the result to the total number of cases from the previous day to forecast the cumulative case. We consider several weight functions 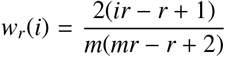 for an *m*-day WMA, where *r* ∈ {1, 2, 4, 10}. The following table shows the different weights for the 4−day WMA.

After comparing the results for the different values of *r*, we notice that the higher the value of *r*, the closer the forecasts to the actual values. Below are the forecasts for the different time intervals using *r* = 10.

Based from the results, the forecasts closest to the actual data among the types of moving average is the 3-day WMA with weight function 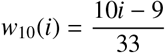.

## 4. Exponential Smoothing

### 4.1. Preliminaries

Exponential smoothing models are commonly used in time series forecasting. These models are simple since the forecasts are generated by obtaining weighted averages of the past observations where the weights exponentially decrease as the observations get older. Exponential smoothing models produce reliably accurate forecasts since they can also capture the trend, seasonality, or a combination of both [25].

It can be observed from Figure 1 that the data has an increasing trend but has no seasonality. Hence, we choose Holt’s linear trend method to obtain 15-day ahead forecasts of the total number of confirmed cases in the Philippines for it allows forecasting with trend.

The Holt’s linear trend method (HLTM) extends the simple exponential smoothing method by using two smoothing equations both of which are dependent on the estimated level and estimated trend of the series at a particular time. The *h*-step ahead forecasting formula under this method is given by the following:

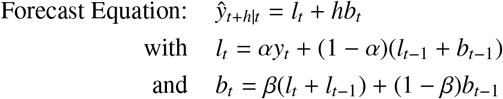

where, at time *t, y*_*t*_ and *ŷ*_*t*+*h*_ represent the actual and the forecast values, respectively, *l*_*t*_ represents the estimated level of the data, and *b*_*t*_ represents the estimated trend of the data. Furthermore, *α* and *β* are the smoothing factors of the level and the trend, respectively, with 0 ≤ *α, β* ≤ 1.

The confidence interval of the forecasts is given by

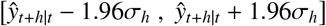

where

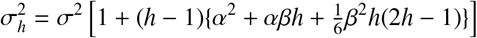

with *σ* as the variance of the forecast errors [25].

### 4.2. Numerical Implementation

We set the initial values of the parameters to be *α* = 0.5 and *β* = 0.5 with *l*_1_ = *y*_1_ and *b*_1_ = *y*_2_ − *y*_1_. We then calculate the 1-day forecasts and the the sum of squared errors (SSE). With this, we obtain the values of *α* and *β* that minimize the SSE subject to the restrictions of *α* and *β*. The values of the parameters that minimize the SSE are *α* = 1 and *β* = 0.5742. These parameters are used to calculate the 15-day ahead forecasts, as well as the forecast confidence interval of the cumulative daily COVID-19 cases in the Philippines. The summary is shown in Table 5.

**Table 5:** 15-day ahead forecasts on the cumulative COVID-19 cases in the Philippines using Holt’s Linear Trend Method.

| Date | Forecast | 95% lower cl | 95% upper cl |
| --- | --- | --- | --- |
| 08/01/2020 | 96966.78 | 96296.45 | 97637.11 |
| 08/02/2020 | 100582.56 | 99332.42 | 101832.7 |
| 08/03/2020 | 104198.33 | 102291.31 | 106105.4 |
| 08/04/2020 | 107814.11 | 105174.54 | 110453.7 |
| 08/05/2020 | 111429.89 | 107987.37 | 114872.4 |
| 08/06/2020 | 115045.67 | 110734.78 | 119356.6 |
| 08/07/2020 | 118661.45 | 113421.01 | 123901.9 |
| 08/08/2020 | 122277.22 | 116049.67 | 128504.8 |
| 08/09/2020 | 125893.00 | 118623.84 | 133162.2 |
| 08/10/2020 | 129508.78 | 121146.17 | 137871.4 |
| 08/11/2020 | 133124.56 | 123618.95 | 142630.2 |
| 08/12/2020 | 136740.33 | 126044.22 | 147436.4 |
| 08/13/2020 | 140356.11 | 128423.79 | 152288.4 |
| 08/14/2020 | 143971.89 | 130759.28 | 157184.5 |
| 08/15/2020 | 147587.67 | 133052.13 | 162123.2 |

As expected from the Holt’s linear trend method, we can see in Figure 4 that the 15-day ahead forecasts exhibit an increasing trend. Moreover, majority of the actual values lie within the prediction interval, suggesting that the method produced fairly reliable forecast values.

**Figure 4:**
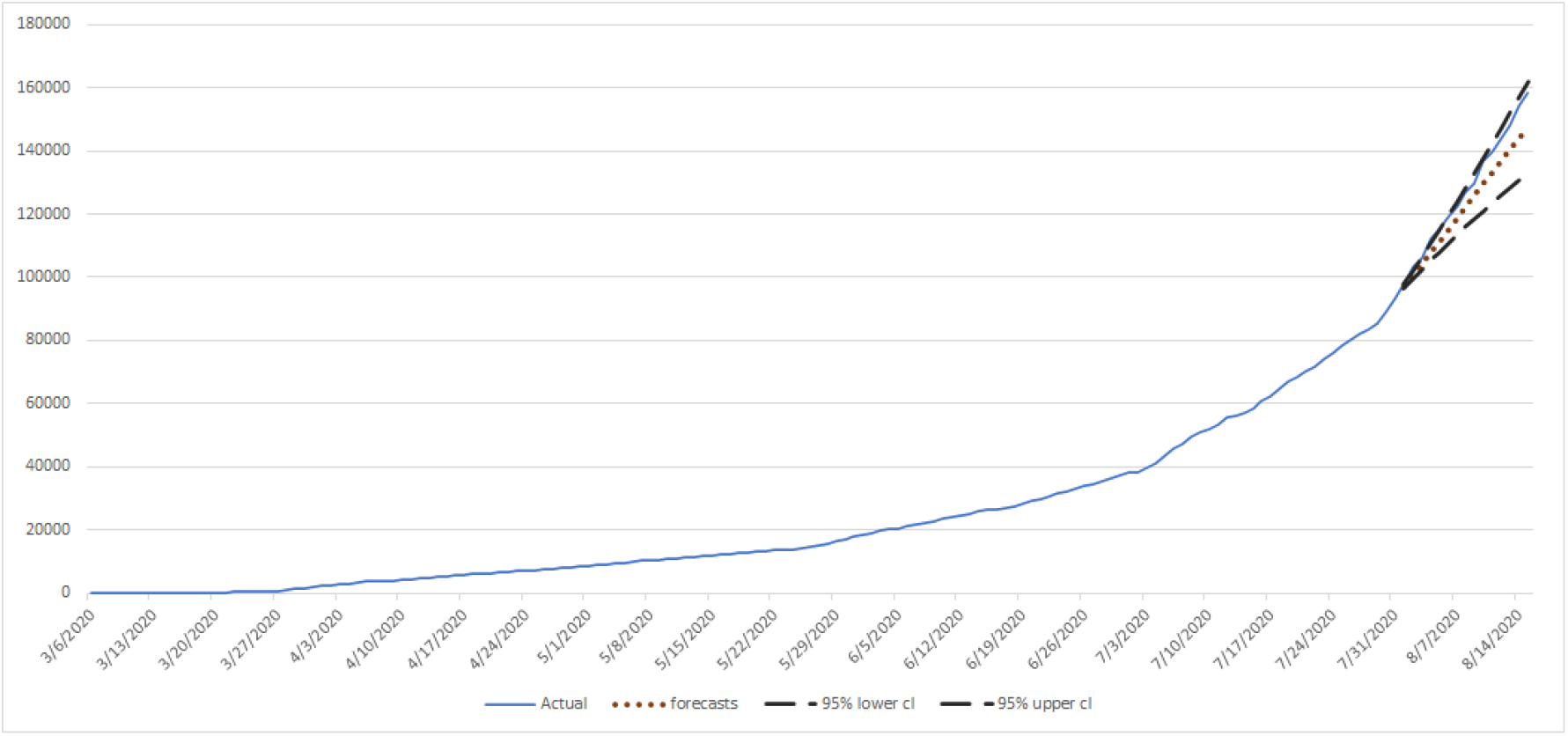
Exponential smoothing using HLTM forecasts vs the actual data.

## 5. SEIR

### 5.1. Preliminaries

#### Model

Differential equation-based models like the SEIR model give information which is useful in controlling different infectious diseases. These models describe disease’s dynamics in macroscopic level [33]. In this study, we also include the Susceptible-Exposed-Infected-Recovered (SEIR) model to describe the dynamics of COVID-19 in the Philippines, taking into account the mortality due to COVID-19 infection. In this model, *S* is the number of susceptible individuals, *E* is the number of exposed individuals, *I* is the number of infected individuals, and *R* is the number of recovered individuals. Susceptible individuals may be in contact with infected individuals and are transferred to the exposed population at rate *β*. A portion *σ* of the exposed individuals will become infected. Infected individuals die at rate *d* or recover from the disease at rate *γ*.

The COVID-19 transmission dynamics is governed by the following set of equations:

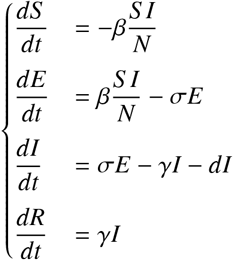

where 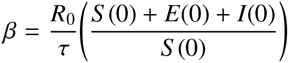 is the transmission coefficient and *N* = *S* + *E* + *I* + *R* represents the total population size [5]. Here, *R*_0_ is the basic reproduction number which is the expected number of secondary infections produced by an infected individual during his or her entire infectious period [16]. Table 6 shows the parameters used in the model and their values obtained from previous studies on COVID-19 while the diagram in Figure 5 describes the inflows and outflows of individuals in each compartment. The parameters near the arrows are rates for which the individuals transfer from one compartment to another.

**Table 6:** Parameters of the COVID-19 transmission model.

| Parameter | Description | Value | Reference |
| --- | --- | --- | --- |
| $R_0$ | Reproduction number | 1.15 | data fitted |
| $\sigma$ | progression rate from $E$ to $I$ | 0.2 | data fitted |
| $\gamma$ | recovery rate | 0.0686 | data fitted |
| $\tau$ | infectious period | 14 | [5] |
| $d$ | death rate due to COVID-19 | 0.03/14 | [6] |

**Figure 5:**
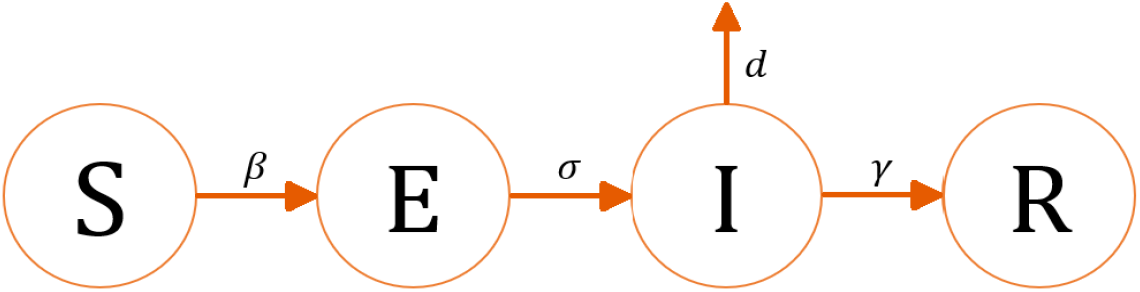
Flow diagram of the COVID-19 transmission model.

#### Sensitivity Analysis

Sensitivity analysis is a method used to identify the effect of each parameter in the model output, which in this case, is the infected population (*I*). In this study, a global sensitivity analysis technique called *partial rank correlation coefficient* (PRCC) analysis is used. To implement PRCC, a stratified random sampling without replacement is used to obtain input parameter values. This method is called Latin Hypercube Sampling (LHS) which is introduced in [30].

In Figure 6, each bar corresponds to a PRCC value at an instance, specifically in days *t* = 67 + 10*k, k* = 0, 1, …, 13. A large absolute PRCC value implies a large correlation of the parameter with the output.The value 1 means a perfect positive linear relationship while −1 means a perfect negative linear relationship. The parameters with high PRCC values (> 0.5 or < −0.5) are *R*_0_, *σ, γ*, and τ. Moreover, *R*_0_ and *σ* have positive PRCC values implying that an increase in the values of these parameters will increase *I*. In contrast, *γ* and *τ* have negative PRCC values which means that a positive change in these parameters will decrease *I*.

**Figure 6:**
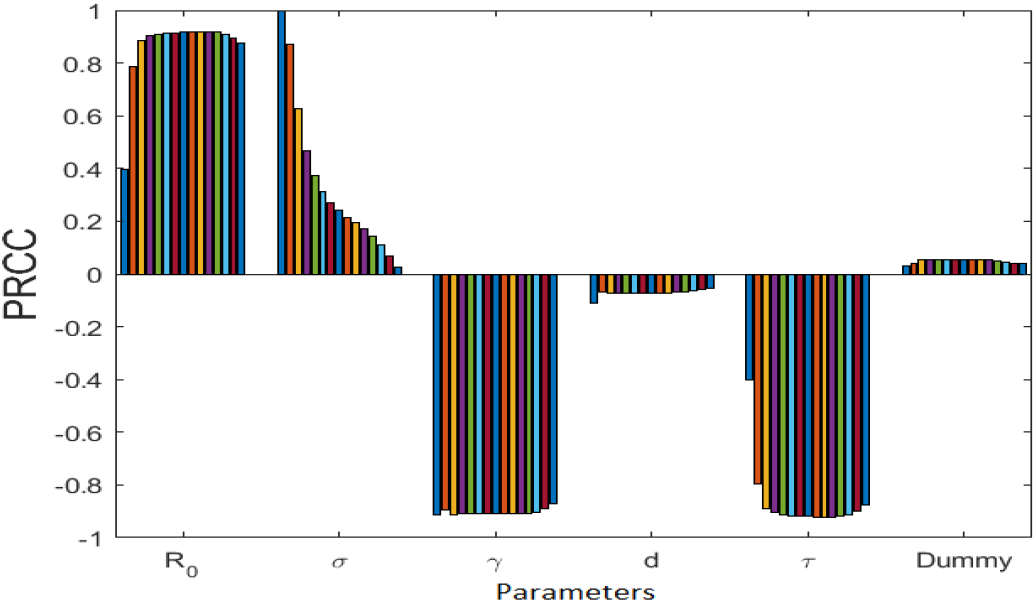
PRCC values showing the sensitivities of the model ouput *I* with respect to the parameters.

#### Parameter Estimation

The output using the SEIR model is made close to the gathered data by estimating some of the parameters for which the model output is sensitive. Reported COVID-19 cases and recoveries in the Philippines are available on the COVID-19 tracker. We use these data sets to estimate the reproduction number (*R*_0_), progression rate from the exposed population to the infected population (*σ*), and the recovery rate (*γ*). The model output was found to have high sensitivity values on these parameters which indicate the parameters’ influential effect to the outcome.We estimate these parameters by minimizing the error between the gathered data and the model output. The estimates are *R*_0_ = 1.5811, *σ* = 0.013, and *γ* = 0.0069. Figure 7 shows the reported data and the corresponding best fit.

**Figure 7:**
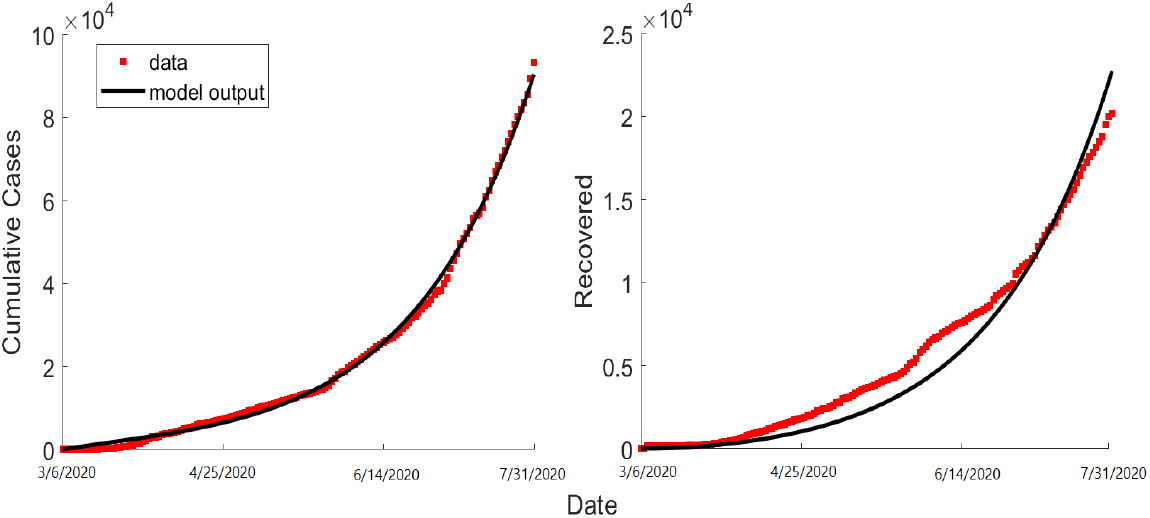
Model fitted curve (black curve) vs actual cumulative cases and recoveries (red)

### 5.2. Numerical Implementation

Using the values obtained in the parameter estimation and the values gathered from other studies on the disease, we forecast the cumulative cases from August 1 - 15 in the country. Table 7 shows the 15-day forecast using the SEIR model and Figure 8 displays the forecast vs actual data.

**Table 7:** 15-day ahead forecasts on the cumulative COVID-19 cases in the Philippines using SEIR.

| Date | Forecast |
| --- | --- |
| 08/01/2020 | 94319.40 |
| 08/02/2020 | 96819.00 |
| 08/03/2020 | 99381.97 |
| 08/04/2020 | 102009.77 |
| 08/05/2020 | 104703.86 |
| 08/06/2020 | 107465.76 |
| 08/07/2020 | 110296.98 |
| 08/08/2020 | 113199.09 |
| 08/09/2020 | 116173.66 |
| 08/10/2020 | 119222.28 |
| 08/11/2020 | 122346.60 |
| 08/12/2020 | 125548.25 |
| 08/13/2020 | 128828.91 |
| 08/14/2020 | 132190.28 |
| 08/15/2020 | 135634.08 |

**Figure 8:**
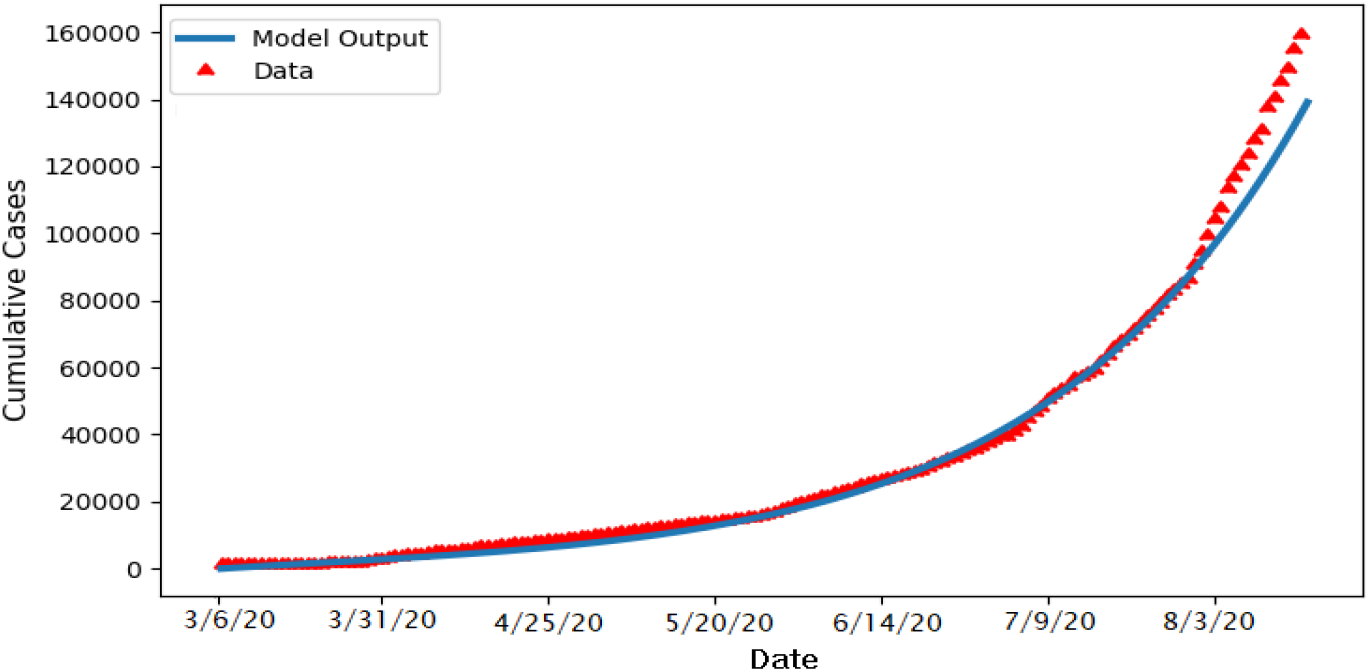
SEIR model forecasts vs the actual data.

Although a similar trend can be perceived in the estimates and the actual data, observe that the SEIR model gave low estimates and the error increases as time passes.This may be attributed to the sudden rise of cases in the month of August as COVID-19 testing in the country improves.

## 6. Ornstein-Uhlenbeck Process

### 6.1. Preliminaries

In this section, we assume that the daily cases denoted by *X*_*t*_, in the discrete sense, is governed by Ornstein-Ulenbeck (OU) process given by the stochastic differential equation

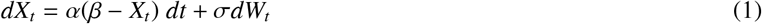

where *α* is the mean reversion, *β* is the drift of the process, *σ* is the volatility, and *W*_*t*_ is a Weiner process. The parameters of *X*_*t*_ are determined using the maximum likelihood estimation. The explicit solution (1) is given by

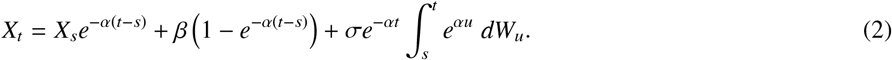

Using Euler-Maruyama approximation, the discrete version of (2) is given by

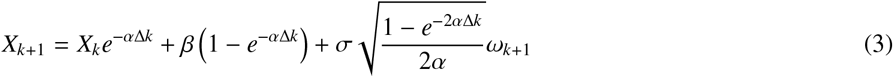

where {*ω*_*k*+1_} is a sequence of IID standard normal random variables.

Let Θ = (*α, β, σ*) be the set of parameters needed with the corresponding likelihood function, *L*(Θ), given by

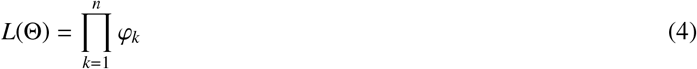

where 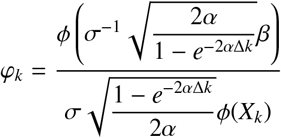 and 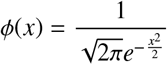.

Notice that the likelihood function is a Radon-Nikodym derivative with respect to the idealized probability measure. Given *n* + 1 observations **X** = {*X*_0_, *X*_1_, …, *X*_*n*_}, the log-likelihood function is

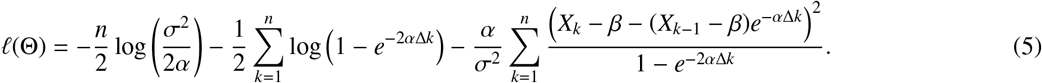

The maximum likelihood estimation requires the gradient of 𝓁 to satisfy the first order conditions, that is,

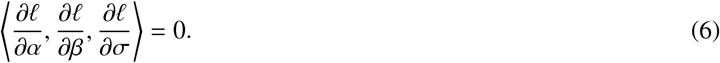

Using the first and second components of (6), we get

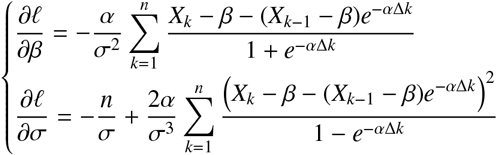

which will give us estimates for *β* and *σ*, given by

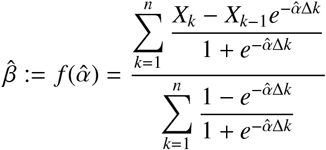

and

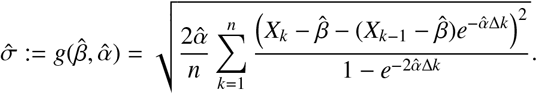

A simpler approach to find an estimate for *α* is to solve the optimization problem

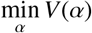

where

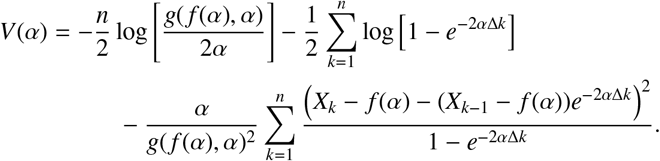

### 6.2. Numerical Implementation

From the data set provided, we determine the parameters of the OU process using the previous subsection’s methodology. Shown in Table 8 are the estimated parameters of the model. Using these parameters, we determine a 15-day forecast depicted in Table 9.

**Table 8:** Estimated parameters of the OU process.

| parameter | Daily cases |
| --- | --- |
| $\alpha$ | 0.05948791 |
| $\beta$ | 1085.746 |
| $\sigma$ | 372.6143 |

**Table 9:** 15-day ahead forecasts on the cumulative COVID-19 cases in the Philippines following OU process.

| Date | Forecast |
| --- | --- |
| 08/01/2020 | 97242.05 |
| 08/02/2020 | 100971.09 |
| 08/03/2020 | 104547.47 |
| 08/04/2020 | 107980.01 |
| 08/05/2020 | 111277.02 |
| 08/06/2020 | 114446.32 |
| 08/07/2020 | 117495.28 |
| 08/08/2020 | 120430.87 |
| 08/09/2020 | 123259.62 |
| 08/10/2020 | 125987.70 |
| 08/11/2020 | 128620.94 |
| 08/12/2020 | 131164.80 |
| 08/13/2020 | 133624.45 |
| 08/14/2020 | 136004.76 |
| 08/15/2020 | 138310.30 |

Shown in Figure 9 is the plot of the forecasted values generated by OU process against the actual recorded values on August 1-15, 2020. If 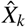 is the forecasted value at time *k*,

**Figure 9:**
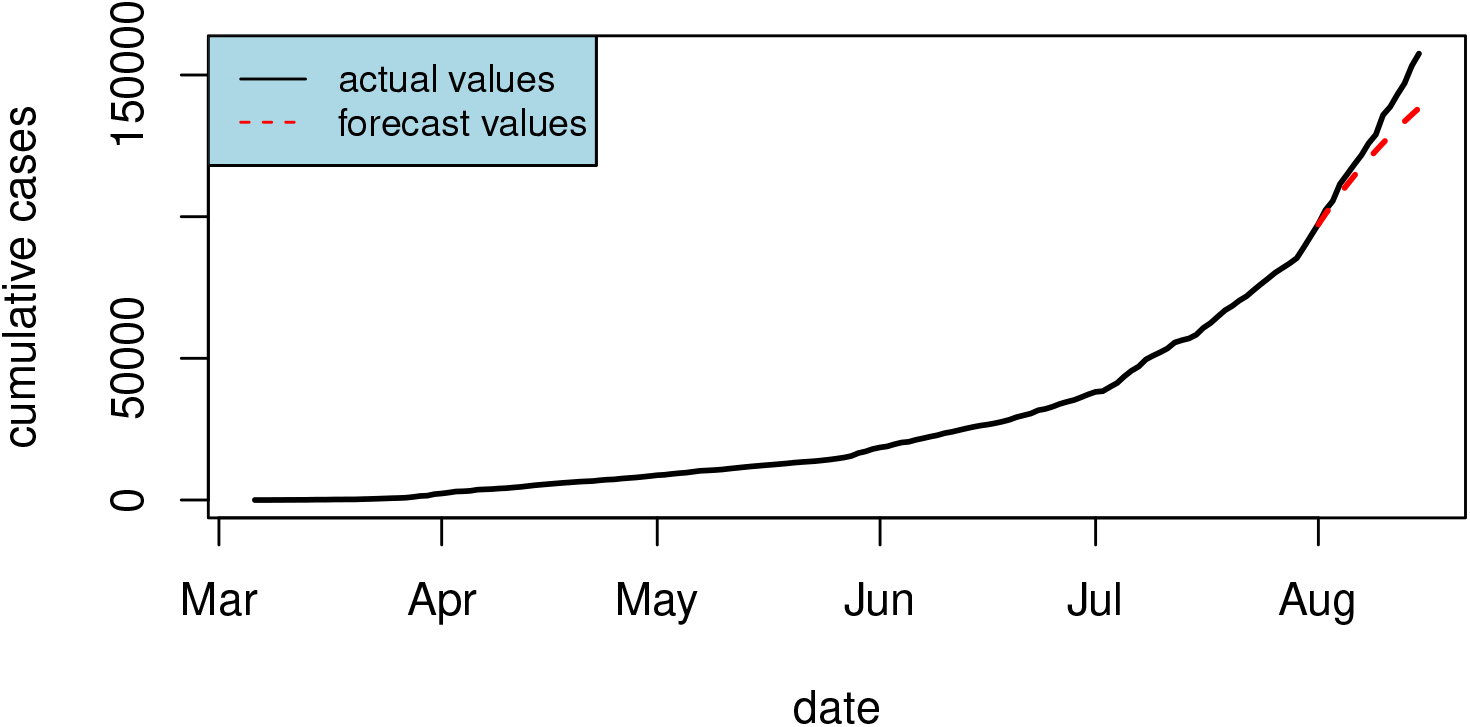
OU process forecast vs the actual data.

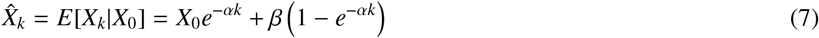

where *X*_0_ is the last actual value required from the forecast, and in our case, the actual value on July 31, 2020. Notice from the plot that the forecast values are slightly bent compared to the actual values, and this is due to the fact that as *k* increases from equation (7), the white noise generated by the actual value from the forecast relatively increases. The noise attributed is 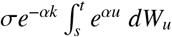 and becomes zero in getting the forecasts.

## 7. ARIMA

Autoregressive Integrated Moving Average (ARIMA) models are widely used in time series modeling and forecasting. This class of models was developed by Box and Jenkins [3] and is intended to describe the process based on historical data. One characteristic of ARIMA models that separate it from other forecasting models is the incorporation of autocorrelation of the data set. This information describes the correlation between observations at different time lags. The general form is given by ARIMA (*p, d, q*), where *p, d*, and *q* refer to the number of autoregressive (AR) parameters, the order of differencing, and the number of moving average (MA) parameters, respectively. The AR component accounts for the pattern between time lags. The MA term measures the adaptation of new forecasts to prior forecast errors. The differencing is performed to remove the trend present in the data set and reduce the non-stationarity [37]. In this section, we forecast the cumulative number of COVID-19 infection cases in the Philippines for August 1-15 using an ARIMA model.

### 7.1. Preliminaries

Due to the increasing trend in the data set as observed in Figure 1, this results in the nonstationarity of the data set. To remove the trend, we perform first-order differencing. Using the Augmented Dickey-Fuller (ADF) test for stationarity, it was found out that the differenced data set is nonstationary at *α* = 0.05. We apply another first-order differencing (equivalently a second-order differencing to the original data set) and resulted in a stationary process using ADF at *α* = 0.05. The resulting stationary process is illustrated in Figure 10.

**Figure 10:**
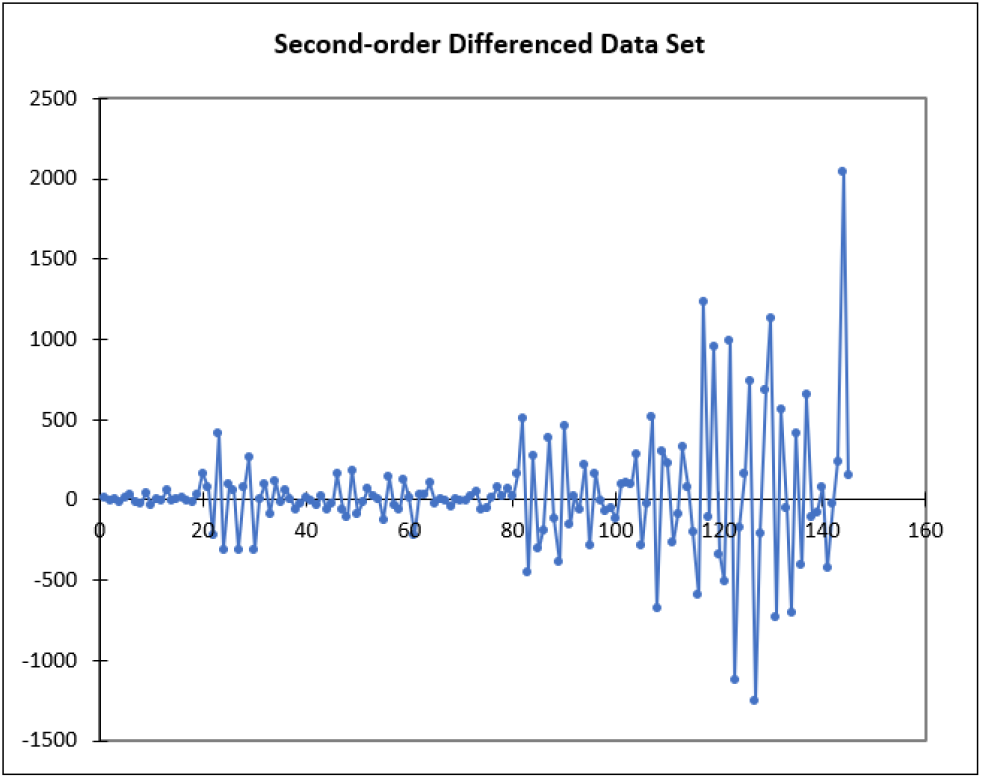
Second-order differenced data on the cumulative cases of infection in the Philippines.

### 7.2. Numerical Implementation

We execute a correlogram analysis on the second-order differenced data set and the autocorrelation (ACF) and partial autocorrelation (PACF) plots are presented in Figure 11. Significant spikes at lag one in the ACF and PACF suggest that an autoregressive (AR) process and a moving average (MA) process at that lag may help explain the data. We examine several models for different values of *p* (order of AR) and *q* (order of MA), with *p* and *q* capped at 5. We compare these models based on the Akaike information criterion (AICc) and the Schwarz’s criterion (SBC). Based on the AICc, the best fit model is ARIMA (5,2,4), while ARIMA (1,2,1) is found to the best fit using SBC. The comparison of the models based on AICc and SBC are shown in Table 10. In Figure 12, ARIMA (1,2,1) and ARIMA (5,2,4) are plotted against the original time series.

**Table 10:** Comparison of the two ARIMA models based on AICc and SBC.

| $p$ | $q$ | AICc | SBC | $p$ | $q$ | AICc | SBC |
| --- | --- | --- | --- | --- | --- | --- | --- |
| **1** | **1** | 1969.901 | **1978.526** | 3 | 4 | 1978.520 | 2000.888 |
| 1 | 2 | 1971.900 | 1983.340 | 3 | 5 | 1967.8840 | 2003.468 |
| 1 | 3 | 1974.060 | 1988.281 | 4 | 1 | 1975.091 | 1992.062 |
| 1 | 4 | 1975.548 | 1992.519 | 4 | 2 | 1977.234 | 1996.920 |
| 1 | 5 | 1976.678 | 1996.364 | 4 | 3 | 1973.700 | 1996.068 |
| 2 | 1 | 1972.674 | 1984.114 | 4 | 4 | 1974.510 | 1999.525 |
| 2 | 2 | 1971.253 | 1985.474 | 4 | 5 | 1979.347 | 2006.973 |
| 2 | 3 | 1976.241 | 1993.211 | 5 | 1 | 1980.538 | 1995.572 |
| 2 | 4 | 1977.894 | 1997.581 | 5 | 2 | 1979.330 | 2001.698 |
| 2 | 5 | 1979.117 | 2002.718 | 5 | 3 | 1981.525 | 2006.540 |
| 3 | 1 | 1972.936 | 1987.157 | **5** | **4** | **1967.334** | 1994.960 |
| 3 | 2 | 1975.193 | 1992.164 | 5 | 5 | 1978.153 | 2008.353 |
| 3 | 3 | 1977.936 | 1997.623 |  |  |  |  |

**Figure 11:**
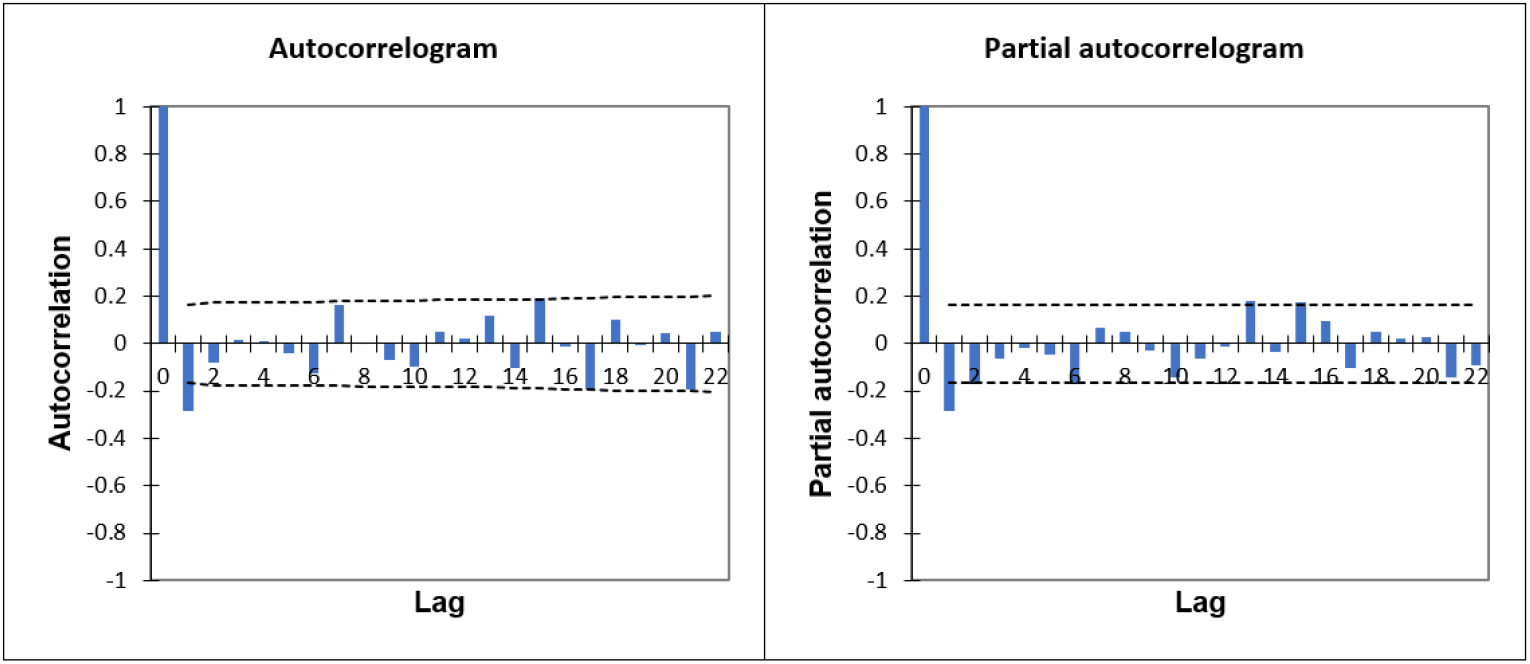
ACF and PACF plots of the second-order differenced data.

**Figure 12:**
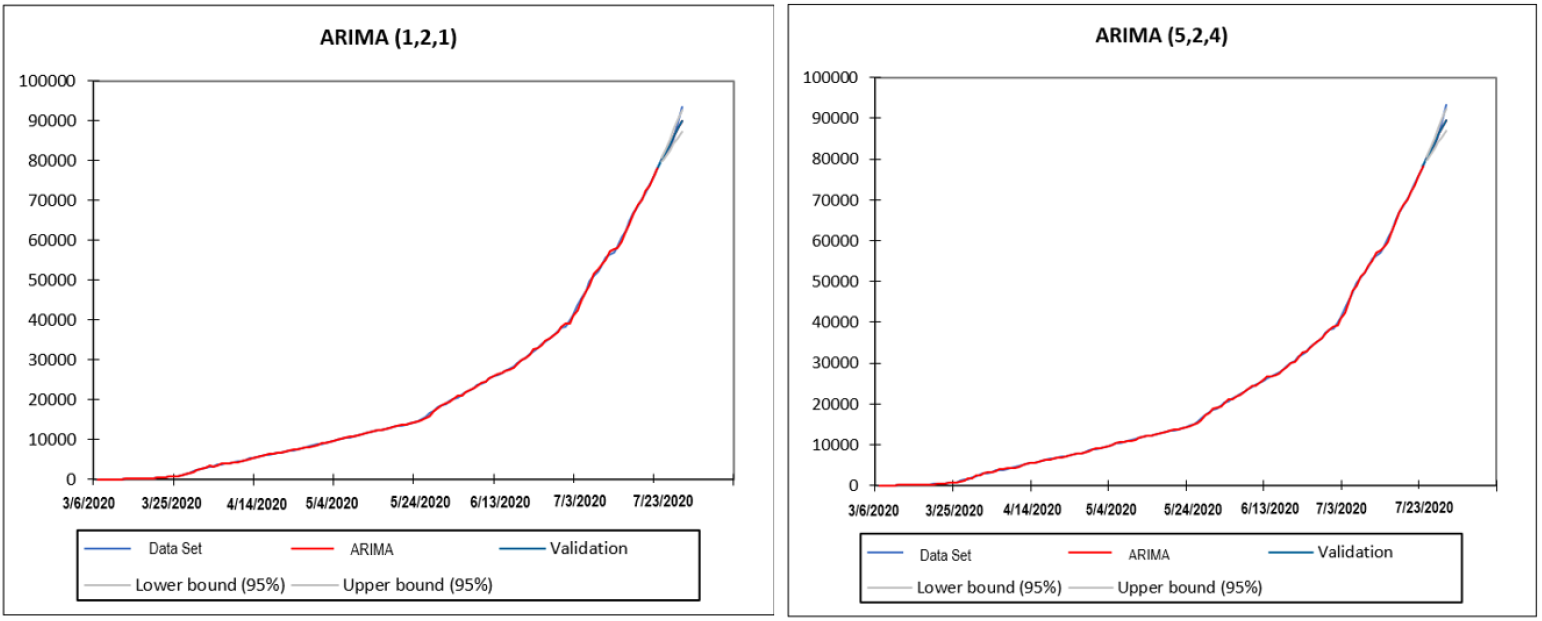
ARIMA(1,2,1) and ARIMA (5,2,4) models vs the actual data.

**Figure 13:**
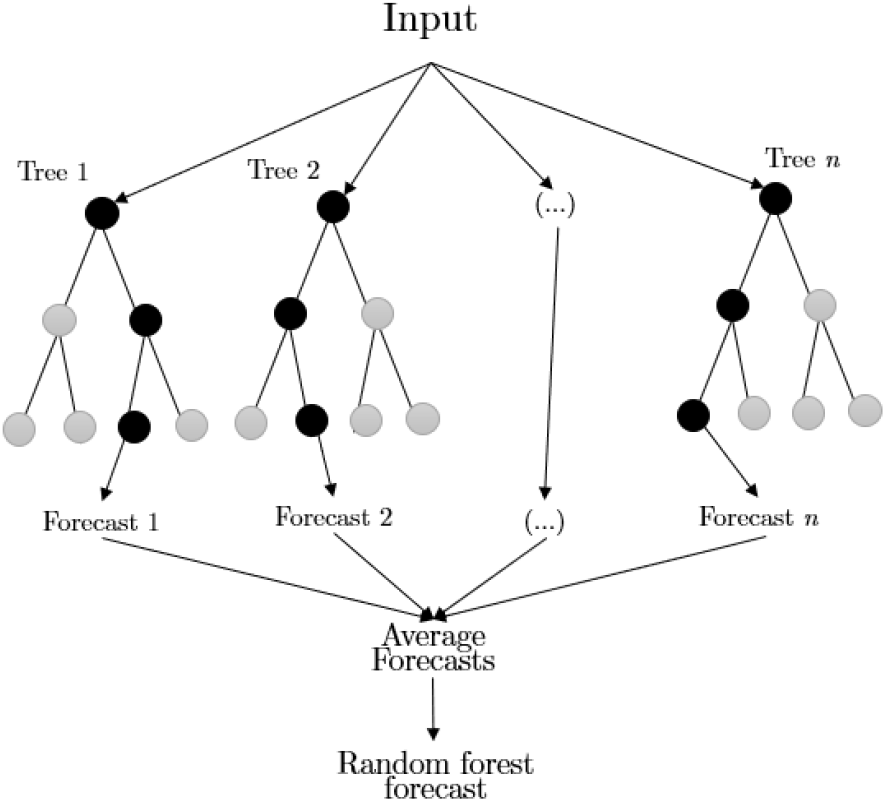
Forecasting process using random forest given *n* number of trees.

The corresponding goodness-of-fit statistics for the two models are summarized in Table 11.

**Table 11:** Goodness-of-fit statistics of the two ARIMA models.

| STATISTICS | ARIMA (1,2,1) | ARIMA (5,2,4) |
| --- | --- | --- |
| SSE | 11073309.7 | 9359918.43 |
| MSE | 79664.0989 | 67337.5426 |
| RMSE | 282.248293 | 259.494783 |
| WN Variance | 79664.0989 | 67337.5426 |
| MAPE(Diff) | 174.524932 | 182.852761 |
| MAPE | 2.68510318 | 2.58111358 |
| -2Log(Like.) | 1963.72275 | 1945.61506 |
| FPE | 80818.651 | 72362.7324 |
| AICc | 1969.90053 | 1967.33381 |
| SBC | 1978.52617 | 1994.9598 |

Using the statistics in Table 11, we perform the Likelihood Ratio Test. At *α* = 0.05, the ARIMA(5,2,4) model does not fit the data significantly better than ARIMA (1,2,1). Therefore, the final ARIMA model is ARIMA(1,2,1) and its parameters are presented in Table 12.

**Table 12:**
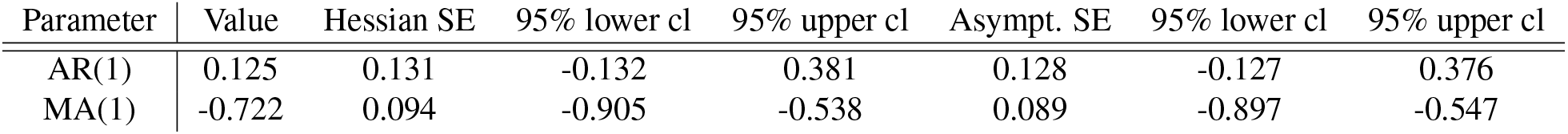
Parameters of ARIMA(1,2,1)

| Parameter | Value | Hessian SE | 95% lower cl | 95% upper cl | Asympt. SE | 95% lower cl | 95% upper cl |
| --- | --- | --- | --- | --- | --- | --- | --- |
| AR(1) | 0.125 | 0.131 | -0.132 | 0.381 | 0.128 | -0.127 | 0.376 |
| MA(1) | -0.722 | 0.094 | -0.905 | -0.538 | 0.089 | -0.897 | -0.547 |

Based on the model, the 15-day forecasts of the daily cumulative cases are presented in Table 13.

**Table 13:** 15-day ahead forecasts on the cumulative COVID-19 cases in the Philippines using ARIMA (1,2,1)

| Date | Forecast |
| --- | --- |
| 08/01/2020 | 97432.88 |
| 08/02/2020 | 101517.1 |
| 08/03/2020 | 105601.6 |
| 08/04/2020 | 109686.2 |
| 08/05/2020 | 113770.8 |
| 08/06/2020 | 117855.3 |
| 08/07/2020 | 121939.9 |
| 08/08/2020 | 126024.5 |
| 08/09/2020 | 130109.1 |
| 08/10/2020 | 134193.6 |
| 08/11/2020 | 138278.2 |
| 08/12/2020 | 142362.8 |
| 08/13/2020 | 146447.3 |
| 08/14/2020 | 150531.9 |
| 08/15/2020 | 154616.5 |

## 8. Random Forest

### 8.1. Preliminaries

Random forest (RF) is a machine learning method for classification and regression problems based from Classification and Regression Tree (CART) and Bagging [31, 18]. Basically, it is a forest made up of a number of decision trees that grow in randomly selected subspaces of the feature space [18, 4]. The idea is to generate a combination of decision trees which will be bootstrapped from the learning sample and from a subset of random features from each chosen node. Every decision tree generated will be built on a subset of learning points and features that were considered from each chosen node to split on. Each tree will be grown to the largest extent possible, and there will be no pruning. After these trees have been fitted through bootstrapping, new forecast will be generated from averaging the forecasts of the trees [18]. An illustration of the process of forecasting in RF is shown below.

Aside from model fitting, RF has been extensively used in various machine learning forecasting studies such as electricity load, employee turnovers, and even on epidemics forecasting [18, 31, 20, 36]. Recently, it has also been used to analyze, predict, and evaluate COVID-19 in India, and COVID-19 patient health [27, 26]. Based from [27] on COVID-19 predictions in India, results from the random forest machine learning algorithm outperformed other machine learning methods. Thus, we implement RF to forecast the COVID-19 cumulative cases of infection in the Philippines.

Using the randomForest package in R [29], we first set the training data and testing data to 148 (number of days from March 6 to July 31) and 15 (number of days to be forecasted), respectively. After which, we fit RF on the training data set with the following parameter inputs:

1. **Training data:** COVID-19 cumulative cases of infection in PH from March 6 to July 31, 2020
2. **Types of random forest:** regression
3. **Number of trees:** 1000
4. **Number of candidate split variables at each split:** 1

After this, we set up a data frame to hold the RF model predictions since we are forecasting cases that haven’t been observed yet. We set up a 97.5% confidence interval for our predictions, and compute for the forecasts (point) with an upper and lower bound, and its standard deviation.

### 8.2. Numerical Implementation

We simulate 15-day ahead forecasts for the RF method. Shown in Table 14 are the results.

**Table 14:** 15-day ahead forecasts on the cumulative COVID-19 cases in the Philippines using random forest machine learning ensemble in R.

| Date | Forecast | 97.5% lower cl | 97.5% upper cl | Standard Deviation |
| --- | --- | --- | --- | --- |
| 8/1/2020 | 86572.08 | 72374.94 | 93351 | 5541.346 |
| 8/2/2020 | 87229.36 | 72699.45 | 93351 | 5542.092 |
| 8/3/2020 | 86722.7 | 72374.94 | 93351 | 5505.979 |
| 8/4/2020 | 86623.09 | 73127.31 | 93351 | 5425.259 |
| 8/5/2020 | 86660.43 | 72374.94 | 93351 | 5543.624 |
| 8/6/2020 | 86623.09 | 73127.31 | 93351 | 5425.259 |
| 8/7/2020 | 86778.93 | 73451.29 | 93351 | 5409.764 |
| 8/8/2020 | 86778.93 | 73451.29 | 93351 | 5409.764 |
| 8/9/2020 | 86711.77 | 72699.45 | 93351 | 5498.981 |
| 8/10/2020 | 86778.93 | 73451.29 | 93351 | 5409.764 |
| 8/11/2020 | 86615.32 | 72814.91 | 93351 | 5442.946 |
| 8/12/2020 | 86615.32 | 72814.91 | 93351 | 5442.946 |
| 8/13/2020 | 86595.98 | 72374.94 | 93351 | 5532.655 |
| 8/14/2020 | 87161.19 | 74044.07 | 93351 | 5708.67 |
| 8/15/2020 | 87161.19 | 74044.07 | 93351 | 5708.67 |

From Figure 14, notice that RF produced underestimates compared to the actual values for the August 1 - 15 COVID-19 infection cases in the Philippines. Some of the initial values were almost captured on the interval, but the further increase of actual COVID-19 cases data in the Philippines was not captured in our result, which showed an almost linear trend based from the plot. Abundance in data is necessary for a better forecast for a machine learning algorithm, which was not evident in this case. Increasing the number of trees did not also differ from the result generated with 1000 trees.

**Figure 14:**
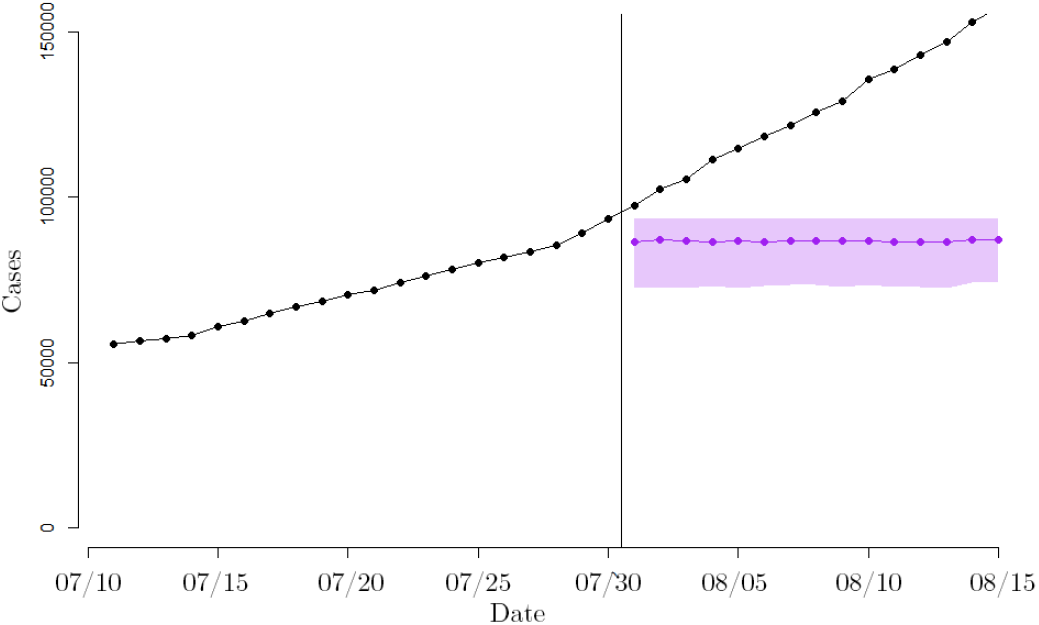
RF forecasts vs the actual data.

## 9. Analysis of Forecast Errors

We plot the 15-day ahead forecasts for August 1 - 15, 2020 from the six models: 3-day WMA, exponential smoothing using HLTM, SEIR model, OU process, ARIMA model, and RF. We then compare them to the actual cumulative cases of infection data in the Philippines.

From Figure 15, notice that all models except RF were able to follow the trend of the actual data. The RF model failed to observe an increasing trend compared to the other models, and was not able to forecast the data well. Of those models that continued to increase, the ARIMA model, 3-day WMA, and HLTM were able to maintain a difference of less than 10,000 cases from the actual data, given the considered forecast period. As the 15th day forecast was approached, the ARIMA(1,2,1) model is closest to the actual values.

**Figure 15:**
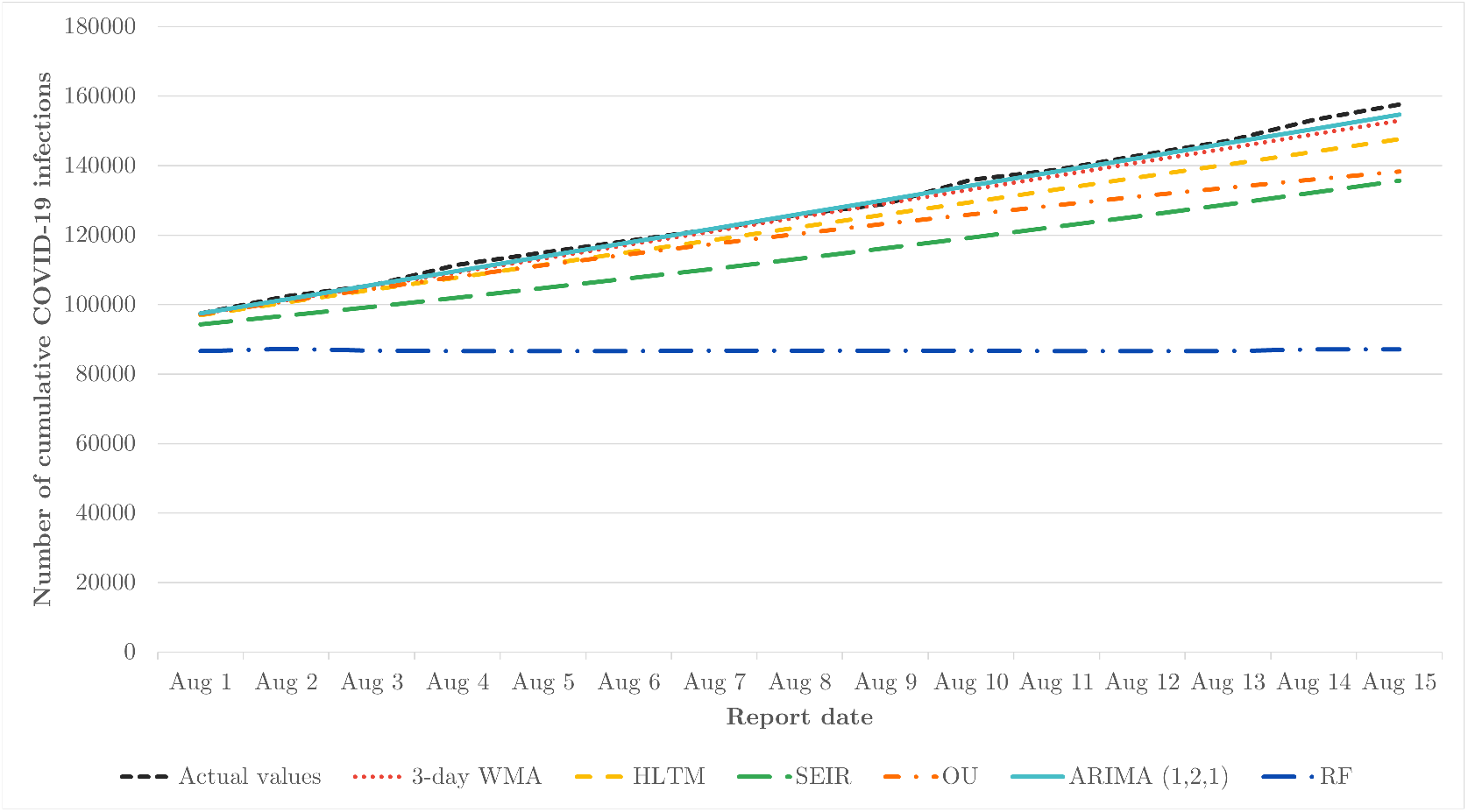
Plot of 15-day-ahead forecast points of the cumulative COVID-19 cases in the Philippines of various mathematical models vs the actual data.

An error analysis of the deviations among the model forecasts of the cumulative cases of infection data in the country is shown in Table 15. The error metrics used are root mean square error (RMSE), absolute mean error (MAE), and relative absolute error (RAE). The RMSE is calculated through 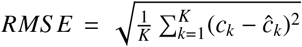, MAE is calculated through 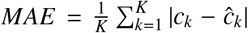, and RAE is calculated through 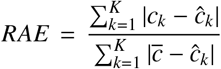, where *c*_*k*_ are the actual values, *ĉ*_*k*_ are the forecasted values, and 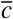 is the mean of the actual values with *K* = 15. From all the error metrics, the ARIMA(1,2,1) model forecasts have the least errors while the random forest forecasts have the highest. Therefore, among the six models used in this study, ARIMA(1,2,1) has the best forecasts of cumulative COVID-19 cases in the Philippines from August 1-15, 2020.

**Table 15:** Error metrics for the forecasted values.

| Error metric | RMSE | MAE | RAE |
| --- | --- | --- | --- |
| 3-day WMA | 2150.65 | 1701.00 | 0.113736 |
| HLTM | 5265.00 | 4536.51 | 0.325803 |
| SEIR | 14036.70 | 12937.74 | 0.856841 |
| OU | 9357.66 | 7389.56 | 0.622168 |
| **ARIMA(1, 2, 1)** | **1330.69** | **1002.01** | **0.065484** |
| RF | 43934.17 | 40038.51 | 1.000000 |

## 10. Summary and Conclusion

In this study, we obtained 15-day ahead forecasts for the daily cumulative COVID-19 cases in the Philippines using mathematical models namely 3-day weighted moving average, exponential smoothing using Holt’s linear trend method, SEIR model, Ornstein-Uhlenbeck process, ARIMA (1,2,1) model, and random forest. We considered the March 6 to July 31 COVID-19 infection data (based from public confirmation date) retrieved from the DOH COVID-19 Data Drop. All models except the random forest produced forecasts that exhibit an increasing trend, with ARIMA (1,2,1) having the closest forecast values to the actual August 1 - 15 data based on the error analysis.

Results from the study can be used by policymakers in determining which forecast method to use/ adapt in their community, especially to those with case behavior similar to the Philippines. These 15-day ahead forecasts provide data-based information for the preparation of the personnel and facilities, and delivery of an effective response. Others may also replicate the methods used and might have a different “best model” for their community. We do note that all models used in the study were based on the retrieved data drop only. Thus, factors that might affect the actual data values such as delay or error in reporting were not considered and might have an effect to the result of this study. A longer amount of data considered might also produce more accurate forecast results. Moreover, we only considered cumulative cases based from the public confirmation date and thus, further studies may consider other data in the data drop, such as cases based on date specimen collection, release of result, date of infection, or even death/ recovery data. We also recommend to explore other models or an ensemble of models, e.g., another machine learning method since the RF model did not do well on our data.

## Data Availability

All data are available upon request to the authors.

